# REACT-1 round 9 interim report: downward trend of SARS-CoV-2 in England in February 2021 but still at high prevalence

**DOI:** 10.1101/2021.02.18.21251973

**Authors:** Steven Riley, Caroline E. Walters, Haowei Wang, Oliver Eales, David Haw, Kylie E. C. Ainslie, Christina Atchison, Claudio Fronterre, Peter J. Diggle, Deborah Ashby, Christl A. Donnelly, Graham Cooke, Wendy Barclay, Helen Ward, Ara Darzi, Paul Elliott

## Abstract

**Background and Methods:** England entered its third national lockdown of the COVID-19 pandemic on 6th January 2021 with the aim of reducing the daily number of deaths and pressure on healthcare services. The real-time assessment of community transmission study (REACT-1) obtains throat and nose swabs from randomly selected people in England in order to describe patterns of SARS-CoV-2 prevalence. Here, we report data from round 9a of REACT-1 for swabs collected between 4th and 13th February 2021.

**Results:** Out of 85,473 tested-swabs, 378 were positive. Overall weighted prevalence of infection in the community in England was 0.51%, a fall of more than two thirds since our last report (round 8) in January 2021 when 1.57% of people tested positive. We estimate a halving time of 14.6 days and a reproduction number R of 0.72, based on the difference in prevalence between the end of round 8 and the beginning of round 9. Although prevalence fell in all nine regions of England over the same period, there was greater uncertainty in the trend for North West, North East, and Yorkshire and The Humber. Prevalence fell substantially across all age groups with highest prevalence among 18- to 24-year olds at 0.89% (0.47%, 1.67%) and those aged 5 to12 years at 0.86% (0.60%, 1.24%). Large household size, living in a deprived neighbourhood, and Asian ethnicity were all associated with increased prevalence. Healthcare and care home workers were more likely to test positive compared to other workers.

**Conclusions:** There is a strong decline in prevalence of SARS-CoV-2 in England among the general population five to six weeks into lockdown, but prevalence remains high: at levels similar to those observed in late September 2020. Also, the number of COVID-19 cases in hospitals is higher than at the peak of the first wave in April 2020. The effects of easing of social distancing when we transition out of lockdown need to be closely monitored to avoid a resurgence in infections and renewed pressure on health services.

## Introduction

The second wave in the COVID-19 pandemic in England started at the beginning of September 2020 [1]. It has been characterized by regional heterogeneity [2] and by alternating periods of relaxed social distancing and prevalence growth [3] followed by increased social distancing and prevalence decline [4]. During December 2020, prevalence of infection increased substantially [5] and led to the highest-to-then levels of hospital admissions due to COVID-19, with southern regions more affected than those in the north [6]. The sudden increase in infections and admissions was partly attributed to a novel variant first detected in the county of Kent in the South East [7]. A legally enforceable third national lockdown started on 6th January 2021 to reduce daily numbers of deaths and to relieve pressure on healthcare systems [8].

The real-time assessment of community transmission study (REACT-1) obtains throat and nose swabs from randomly selected people in England in order to describe patterns of SARS-CoV-2 prevalence [9]. Swabs were collected for round 8 of the study mainly between 6th and 22nd January 2021 and showed a plateau in prevalence for the first 10 days of the period before a fall in the final seven days [10]. Similar patterns were observed in data from the Office for National Statistics Coronavirus Infection Survey [11] and in the proportion of people testing positive among those receiving a routine PCR test in England [6].

Round 9a of REACT-1 commenced self-administered swab-collection on 4th February 2021 and we report here results from swabs collected up to and including 13th February.

## Results

In round 9a, we found 378 positives from 85,473 swabs giving an unweighted prevalence of 0.44% (95% CI, 0.40%, 0.49%) and a weighted prevalence of 0.51% (0.45%, 0.59%) (Table 1). These findings describe a greater than two thirds decrease in weighted prevalence from 1.57% (1.49%, 1.66%) observed during round 8.

**Table 1.**
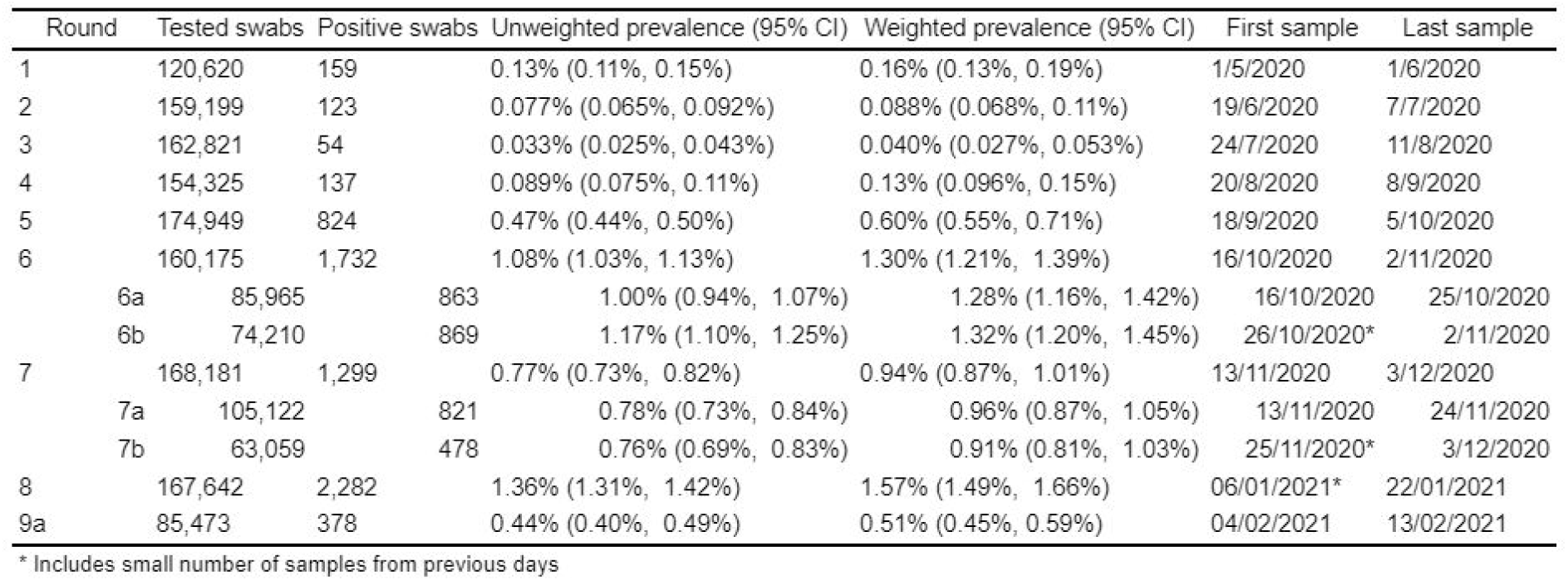
Unweighted and weighted prevalence of swab-positivity across nine rounds of REACT-1.

Fitting a P-spline shows that prevalence fell during the second half of round 8 and continued downwards through round 9a (Figure 1). For the purposes of estimating R, we define round 8b from 16th to 22nd January 2021. Using a constant growth rate model (Table 2), across rounds 8b and 9a, we estimated a halving time of 14.6 (12.7, 17.1) days corresponding to an R of 0.72 (0.69, 0.76). As a sensitivity analysis, we also estimated R across round 8 as a whole through to rounds 9a, giving a halving time of 20.4 (18.6, 22.6) days corresponding to R of 0.80 (0.78, 0.82).

**Table 2.** Estimates of national growth rates, doubling times and reproduction numbers for rounds 8b and 9a, and for rounds 8 and 9a.

| Round | Outcome | Growth rate | R | Probability R>1 | Growth(+)/Decay(-) rate |
| --- | --- | --- | --- | --- | --- |
| 8b and 9a | All positives | -0.048 ( -0.054 , -0.041 ) | 0.72 ( 0.69 , 0.76 ) | <0.01 | -14.6 ( -12.7 , -17.1 ) |
|  | Non-symptomatics | -0.039 ( -0.051 , -0.026 ) | 0.77 ( 0.71 , 0.84 ) | <0.01 | -17.9 ( -13.6 , -26.4 ) |
|  | Positive for both E and N genes | -0.053 ( -0.062 , -0.045 ) | 0.69 ( 0.65 , 0.74 ) | <0.01 | -13.0 ( -11.2 , -15.5 ) |
|  | Positive for both E and N genes or positive only for N gene with CT 35 or less | -0.048 ( -0.056 , -0.040 ) | 0.72 ( 0.68 , 0.76 ) | <0.01 | -14.4 ( -12.5 , -17.1 ) |
| 8 and 9a | All positives | -0.034 ( -0.037 , -0.031 ) | 0.80 ( 0.78 , 0.82 ) | <0.01 | -20.4 ( -18.6 , -22.6 ) |
|  | Non-symptomatics | -0.033 ( -0.039 , -0.027 ) | 0.80 ( 0.77 , 0.84 ) | <0.01 | -21.1 ( -18.0 , -25.5 ) |
|  | Positive for both E and N genes | -0.037 ( -0.042 , -0.033 ) | 0.78 ( 0.75 , 0.80 ) | <0.01 | -18.6 ( -16.6 , -20.9 ) |
|  | Positive for both E and N genes or positive only for N gene with CT 35 or less | -0.035 ( -0.039 , -0.032 ) | 0.79 ( 0.77 , 0.81 ) | <0.01 | -19.6 ( -17.7 , -21.7 ) |

**Figure 1.**
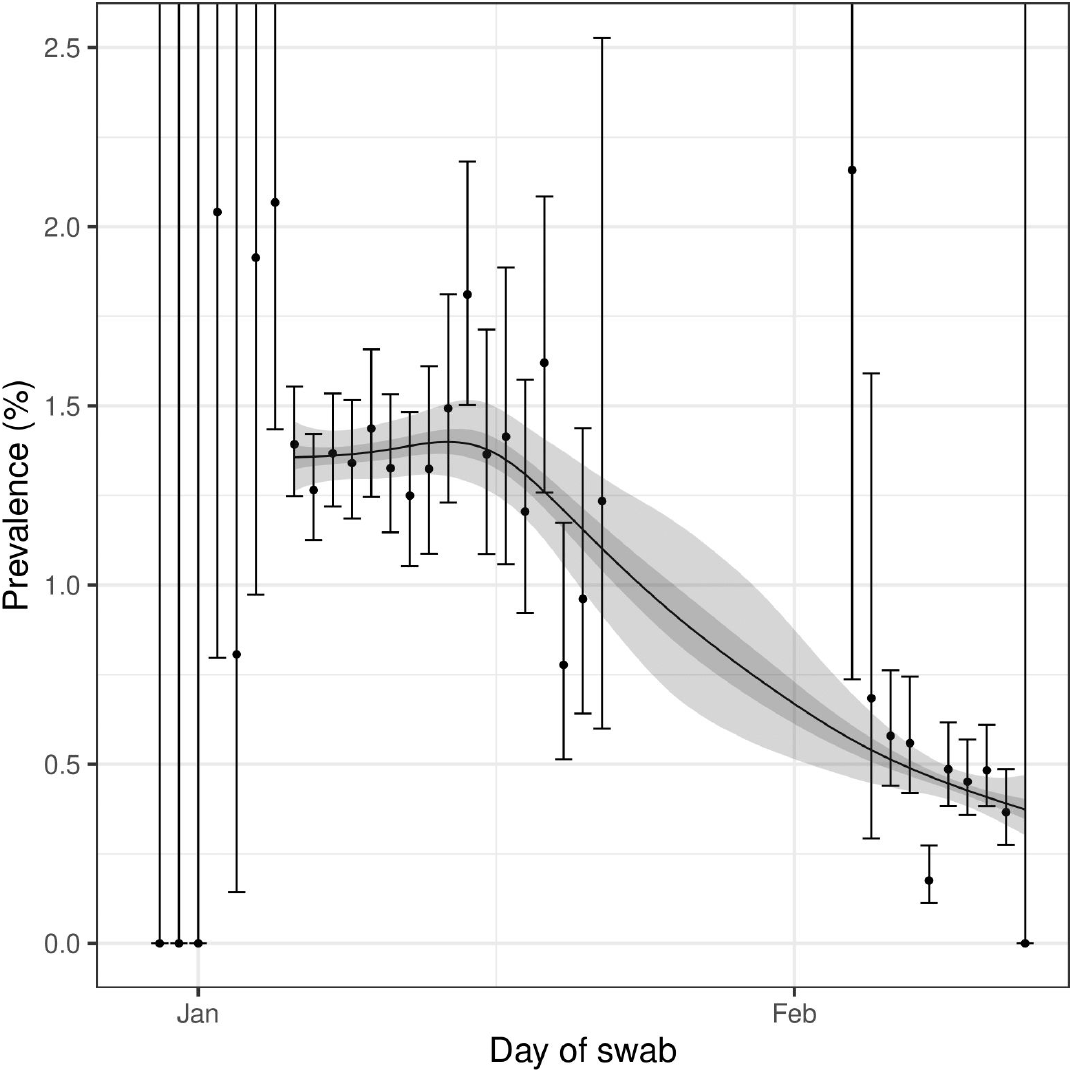
Prevalence of national swab-positivity for England estimated using a p-spline for all nine rounds with central 50% (dark grey) and 95% (light grey) posterior credible intervals. Shown here only for the period of round 8 to round 9a. Vertical lines show the 95% confidence intervals for each data point, note that for some days with small numbers of samples the confidence interval extends beyond the limit of the graph.

Between rounds 8 and 9a, there were falls in weighted regional prevalence (Table 3, Figure 2) with regional prevalence now highest in the North West at 0.91% (0.67%, 1.22%), and North East at 0.82% (0.53%, 1.26%). These falls were substantial in the following regions: London from 2.83% (2.53%, 3.16%) to 0.54% (0.37%, 0.79%), West Midlands from 1.66%(1.32%, 2.10%) to 0.33% (0.22%, 0.52%), East of England from 1.78% (1.57%, 2.02%) to 0.54% (0.39%, 0.75%), South East from 1.61% (1.46%, 1.77%) to 0.30% (0.22%, 0.41%) and East Midlands from 1.16% (0.99%, 1.36%) to 0.51% (0.38%, 0.67%). There was a smaller apparent fall in Yorkshire and The Humber. Regional P-splines showed similar trends (Figure 3) which were largely consistent with trends in proportion of people testing positive through the national surveillance program (Pillar 1 and 2) (Figure 4, Figure 5) [6].

**Table 3.**
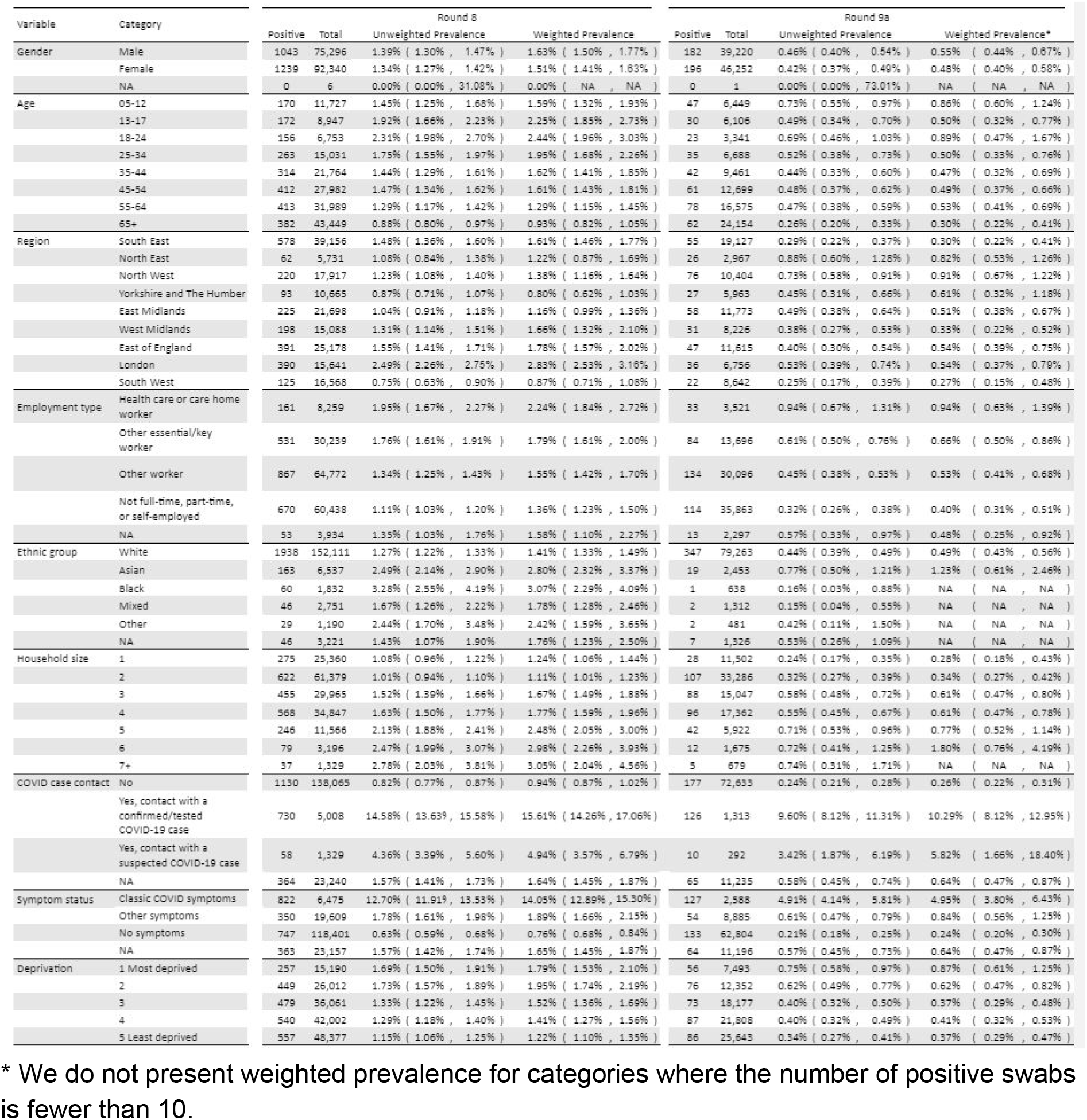
Unweighted and weighted prevalence of swab-positivity for rounds 8 and 9a.

**Figure 2.**
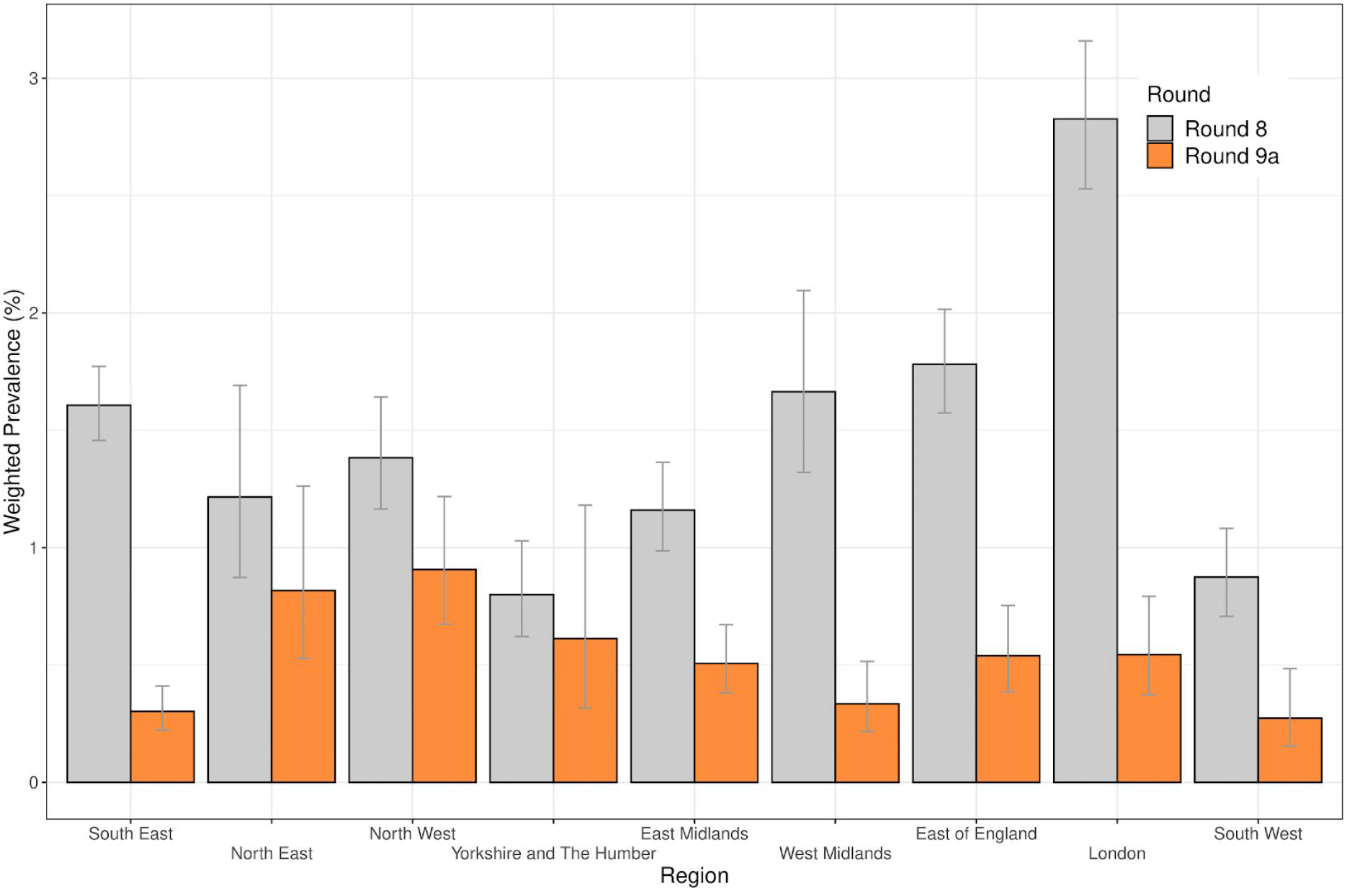
Weighted prevalence of swab-positivity by region for rounds 8 and 9a. Bars show 95% confidence intervals.

**Figure 3.**
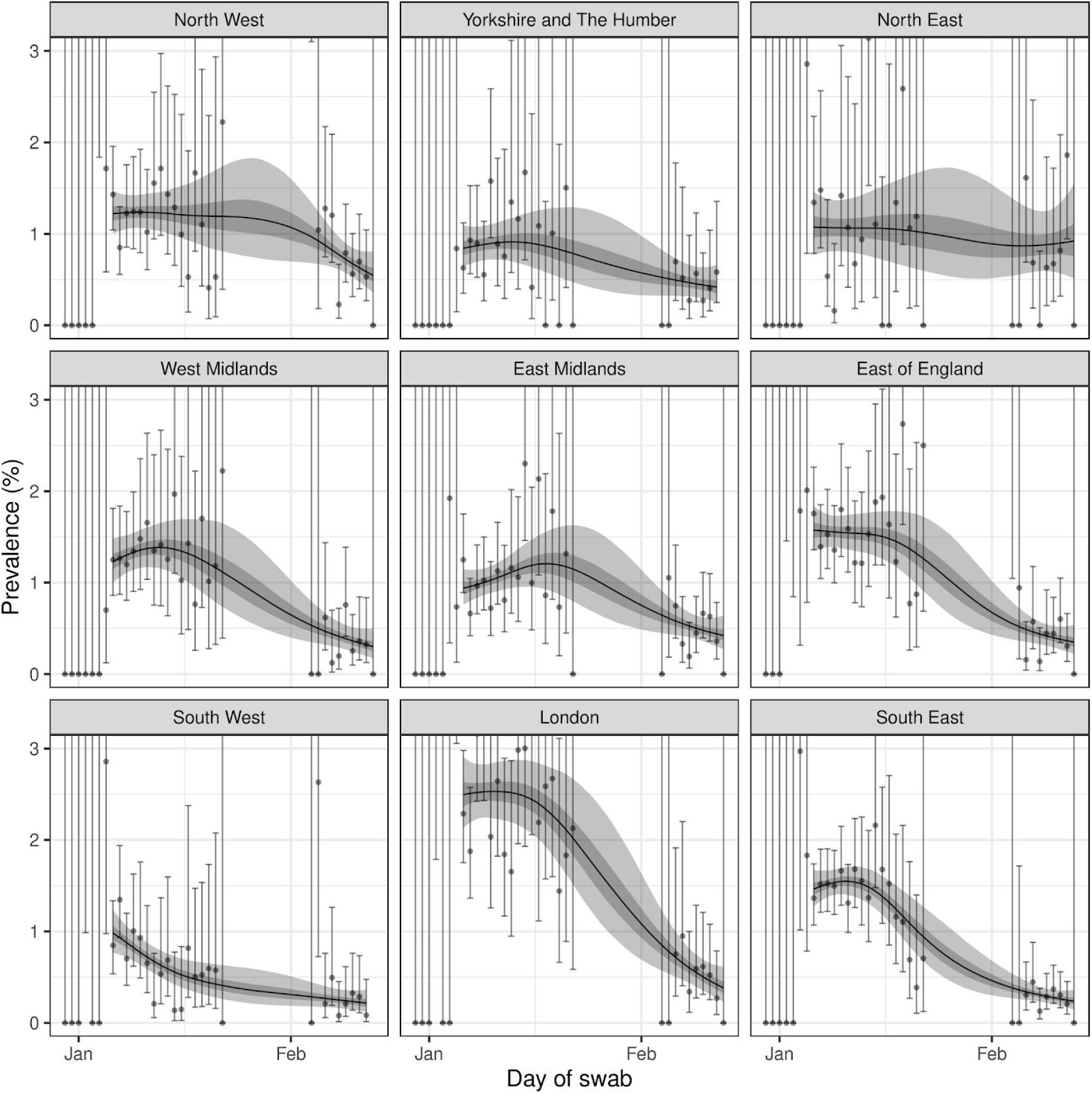
Prevalence of swab-positivity estimated using a P-spline (with constant second-order random walk prior) for each region of England separately. Each model was fit to all nine rounds but is only shown here for the period of round 8 to round 9a. Central 50% (dark grey) and 95% (light grey) posterior credible intervals are also shown. Vertical lines show the 95% confidence intervals for each data point, note that for some days with small numbers of samples the confidence interval extends beyond the limit of the graph.

**Figure 4.**
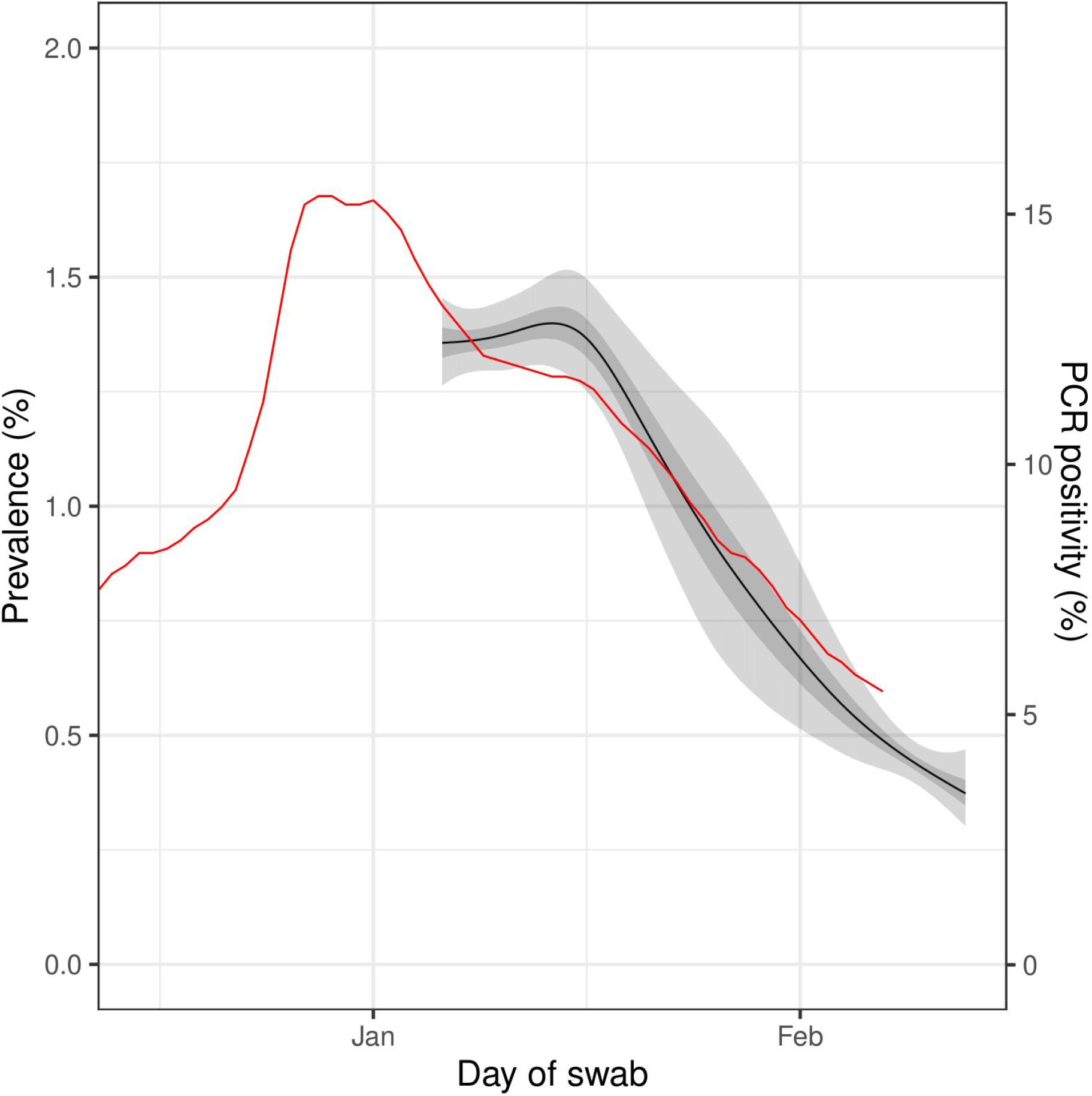
Comparison of REACT-1 estimated prevalence using a P-spline with central 50% (dark grey, left axis) and 95% (light grey, left axis) posterior credible intervals and routine PCR positivity for England (red, right axis) averaged over 7 days, plotted at the midpoint of the interval.

**Figure 5.**
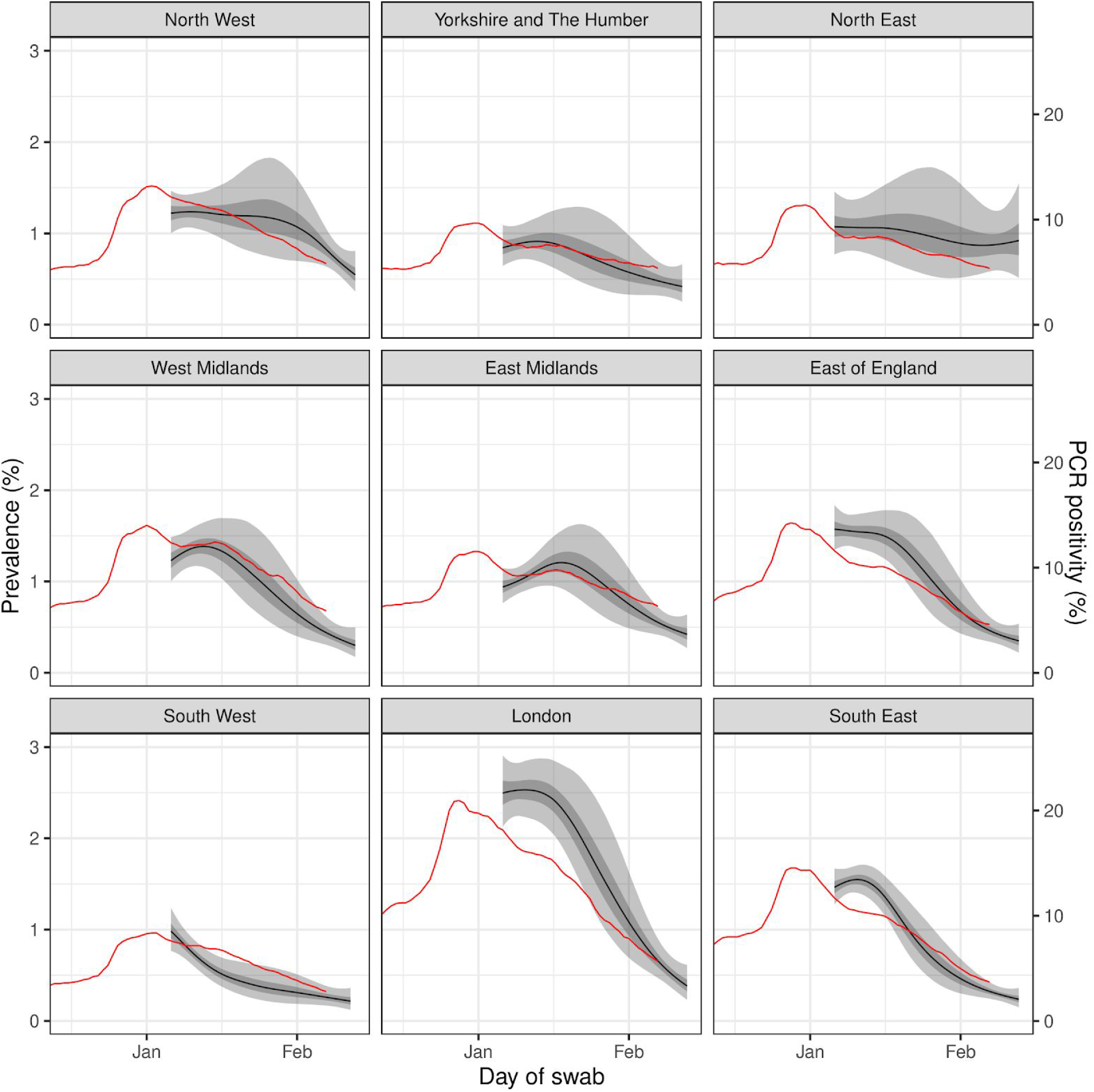
Regional comparison of REACT-1 estimated prevalence using a P-spline with central 50% (dark grey, left axis) and 95% (light grey, left axis) posterior credible intervals and routine PCR positivity for England (red, right axis) averaged over 7 days, plotted at the midpoint of the interval.

Falls in regional prevalence were reflected in regional R estimates (Table 4), which, for the period from round 8b to 9a, were robustly less than one everywhere other than North East, North West, and Yorkshire and The Humber. As a sensitivity analysis, for the period from round 8 to 9a, regional Rs were robustly less than one everywhere other than the North East. Sub-regional patterns highlight the near-universal drop in prevalence across England, but there was suggestion of elevated prevalence in the Greater Manchester / Lancashire area (Figure 6).

**Table 4.**
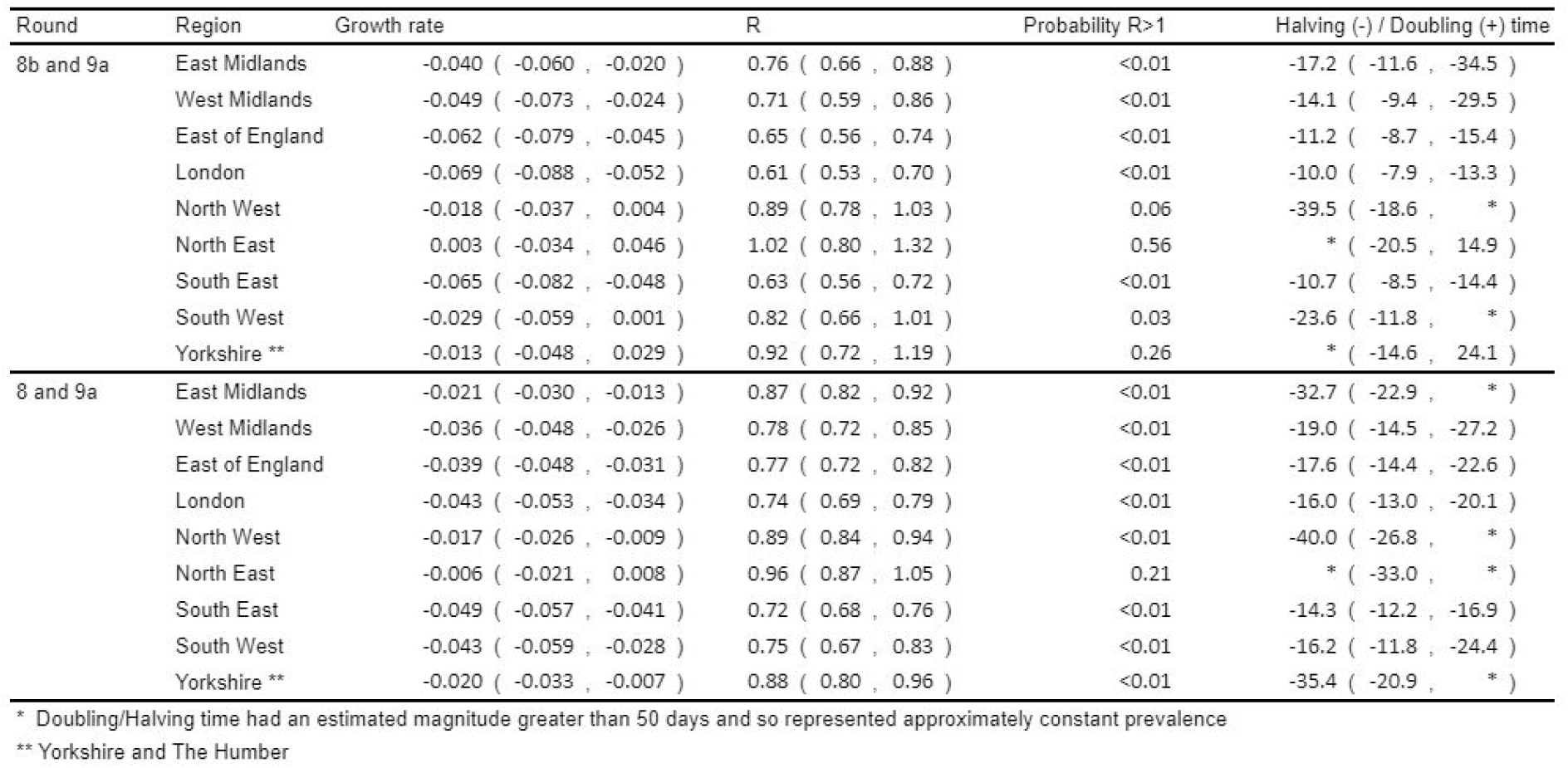
Estimates of regional growth rates, doubling times and reproduction numbers for rounds 8b and 9a, and for rounds 8 and 9a.

**Figure 6.**
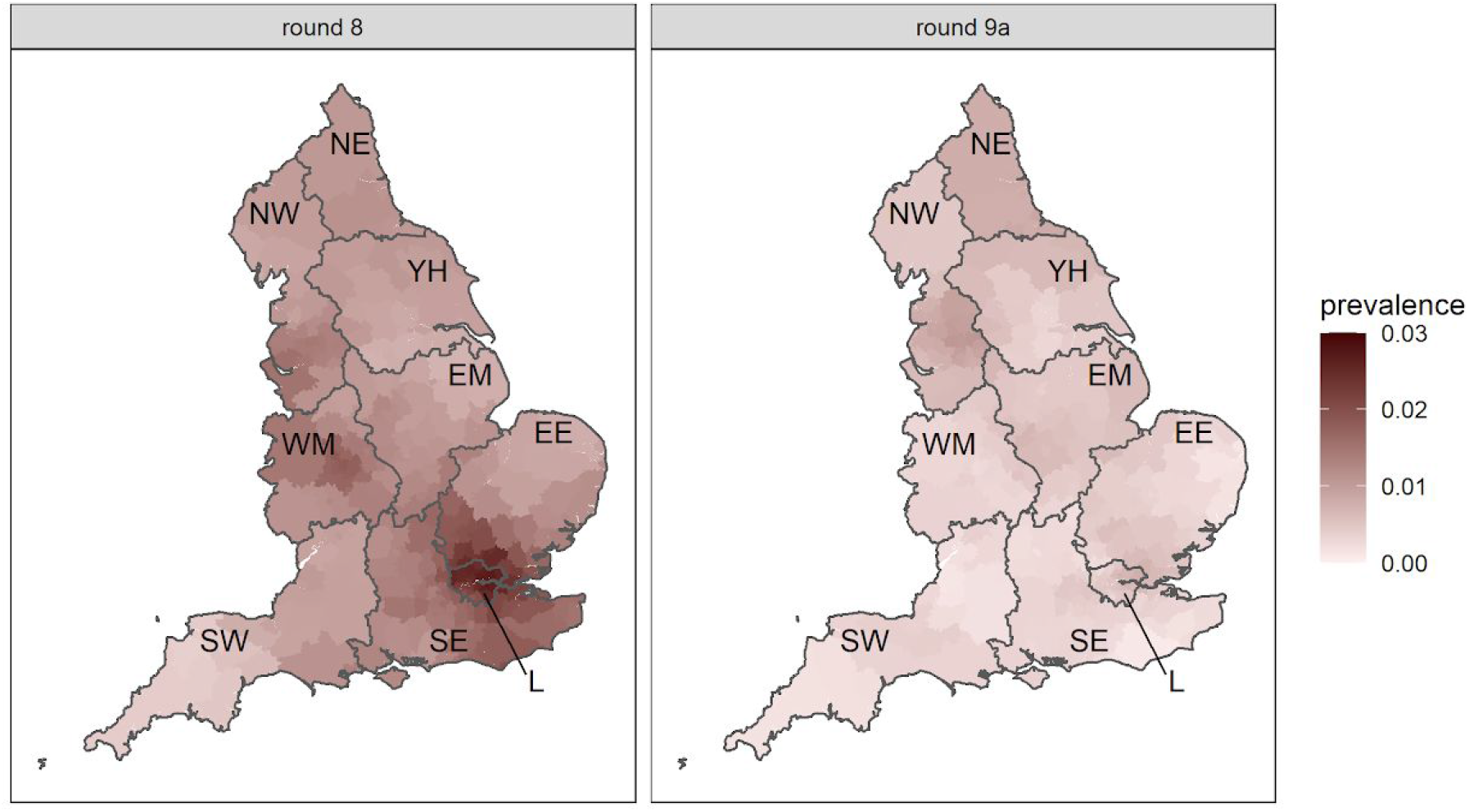
Neighbourhood prevalence of swab-positivity for rounds 8 and 9a. Neighbourhood prevalence calculated from nearest neighbours (the median number of neighbours within 30km in the study). Average neighbourhood prevalence displayed for individual lower-tier local authorities. Regions: NE = North East, NW = North West, YH = Yorkshire and The Humber, EM = East Midlands, WM = West Midlands, EE = East of England, L = London, SE = South East, SW = South West.

There have also been substantial falls in prevalence across all age groups between rounds 8 and 9a (Table 3, Figure 7). Highest weighted prevalence is now among 18 to 24 year olds at 0.89% (0.47%, 1.67%) and ages 5 to 12 years at 0.86% (0.60%, 1.24%). Lowest prevalence is among those 65 years and older at 0.30% (0.22%, 0.41%).

**Figure 7.**
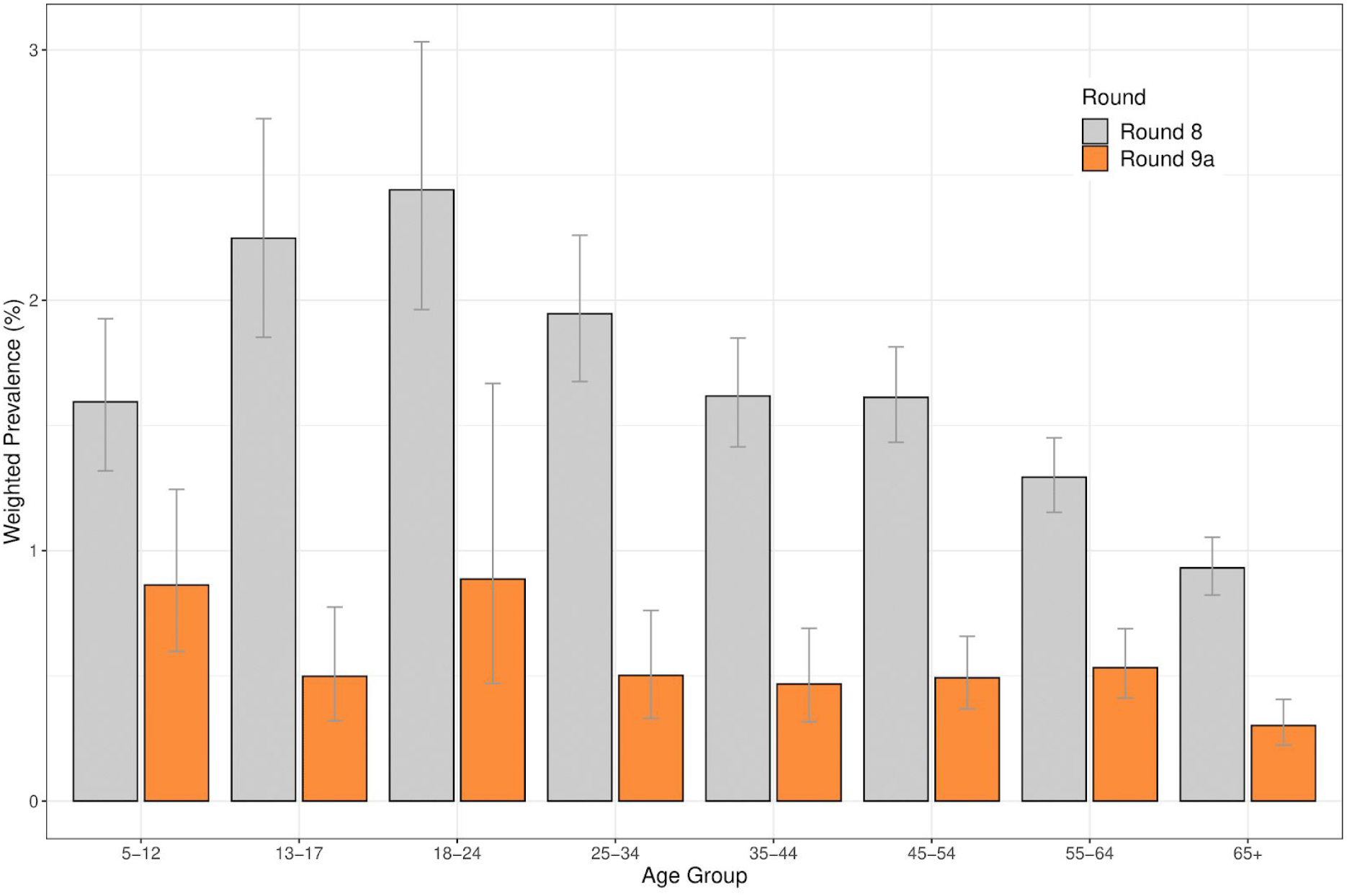
Weighted prevalence of swab-positivity by age groups for rounds 8 and 9a. Bars show 95% confidence intervals.

Patterns of higher prevalence of swab-positivity previously reported [10] were maintained in round 9a among: people of Asian ethnicity compared to white people, healthcare or care home workers versus other workers, people living in most compared to least deprived neighbourhoods and large households versus single person households, although when adjusting for other variables in a regression model, the evidence for increased risk for Asian ethnicity was less certain (Table 5, Figure 8). However we note that the point estimate for the odds ratio for Asian ethnicity in rounds 8 and 9a were similar.

**Table 5.** Jointly adjusted odds ratios for swab-positivity by: sex, age, region, key worker status, ethnicity, household size and index of area deprivation.

| Variable | Category | 8 | 9a |
| --- | --- | --- | --- |
| Sex | Male | Ref | Ref |
|  | Female | 0.95 [0.87,1.03] | 0.83 [0.67,1.03] |
| Age Group | 5-12 | 0.94 [0.78,1.15] | 1.61 [1.04,2.49] |
|  | 13-17 | 1.70 [1.36,2.12] | 1.21 [0.68,2.14] |
|  | 18-24 | 1.71 [1.40,2.09] | 1.75 [1.04,2.93] |
|  | 25-34 | 1.30 [1.10,1.54] | 1.29 [0.82,2.03] |
|  | 35-44 | Ref | Ref |
|  | 45-54 | 1.15 [0.99,1.34] | 1.22 [0.82,1.83] |
|  | 55-64 | 1.19 [1.02,1.39] | 1.54 [1.03,2.30] |
|  | 65+ | 1.02 [0.85,1.22] | 1.14 [0.71,1.82] |
| Region | North East | 0.70 [0.54,0.92] | 2.61 [1.59,4.30] |
|  | North West | 0.78 [0.66,0.92] | 2.31 [1.60,3.34] |
|  | Yorkshire | 0.60 [0.48,0.75] | 1.48 [0.91,2.39] |
|  | East Midlands | 0.69 [0.59,0.81] | 1.67 [1.13,2.45] |
|  | West Midlands | 0.86 [0.73,1.02] | 1.31 [0.83,2.05] |
|  | East of England | 1.01 [0.89,1.16] | 1.37 [0.91,2.05] |
|  | London | 1.47 [1.27,1.69] | 1.83 [1.18,2.84] |
|  | South East | Ref | Ref |
|  | South West | 0.51 [0.42,0.62] | 0.87 [0.52,1.45] |
| Key Worker Status | HCW/CHW | 1.48 [1.25,1.77] | 2.00 [1.35,2.95] |
|  | Key worker (other) | 1.35 [1.20,1.51] | 1.22 [0.92,1.61] |
|  | Other worker | Ref | Ref |
|  | Not FT, PT, SE | 0.90 [0.80,1.02] | 0.78 [0.59,1.04] |
| Ethnicity | Asian | 1.35 [1.14,1.61] | 1.40 [0.86,2.30] |
|  | Black | 1.63 [1.24,2.14] | 0.25 [0.03,1.78] |
|  | Mixed | 1.03 [0.77,1.39] | 0.28 [0.07,1.13] |
|  | Other | 1.39 [0.96,2.03] | 0.39 [0.05,2.77] |
|  | White | Ref | Ref |
| Household Size | 1-2 People | Ref | Ref |
|  | 3-5 People | 1.37 [1.24,1.52] | 1.81 [1.38,2.36] |
|  | 6+ People | 2.02 [1.64,2.50] | 2.02 [1.14,3.59] |
| Deprivation Index Quintile | 1 - Most Deprived | Ref | Ref |
|  | 2 | 1.03 [0.88,1.21] | 0.88 [0.62,1.27] |
|  | 3 | 0.82 [0.70,0.97] | 0.64 [0.44,0.92] |
|  | 4 | 0.82 [0.70,0.96] | 0.64 [0.45,0.90] |
|  | 5 - Least Deprived | 0.72 [0.61,0.84] | 0.56 [0.39,0.80] |

**Figure 8.**
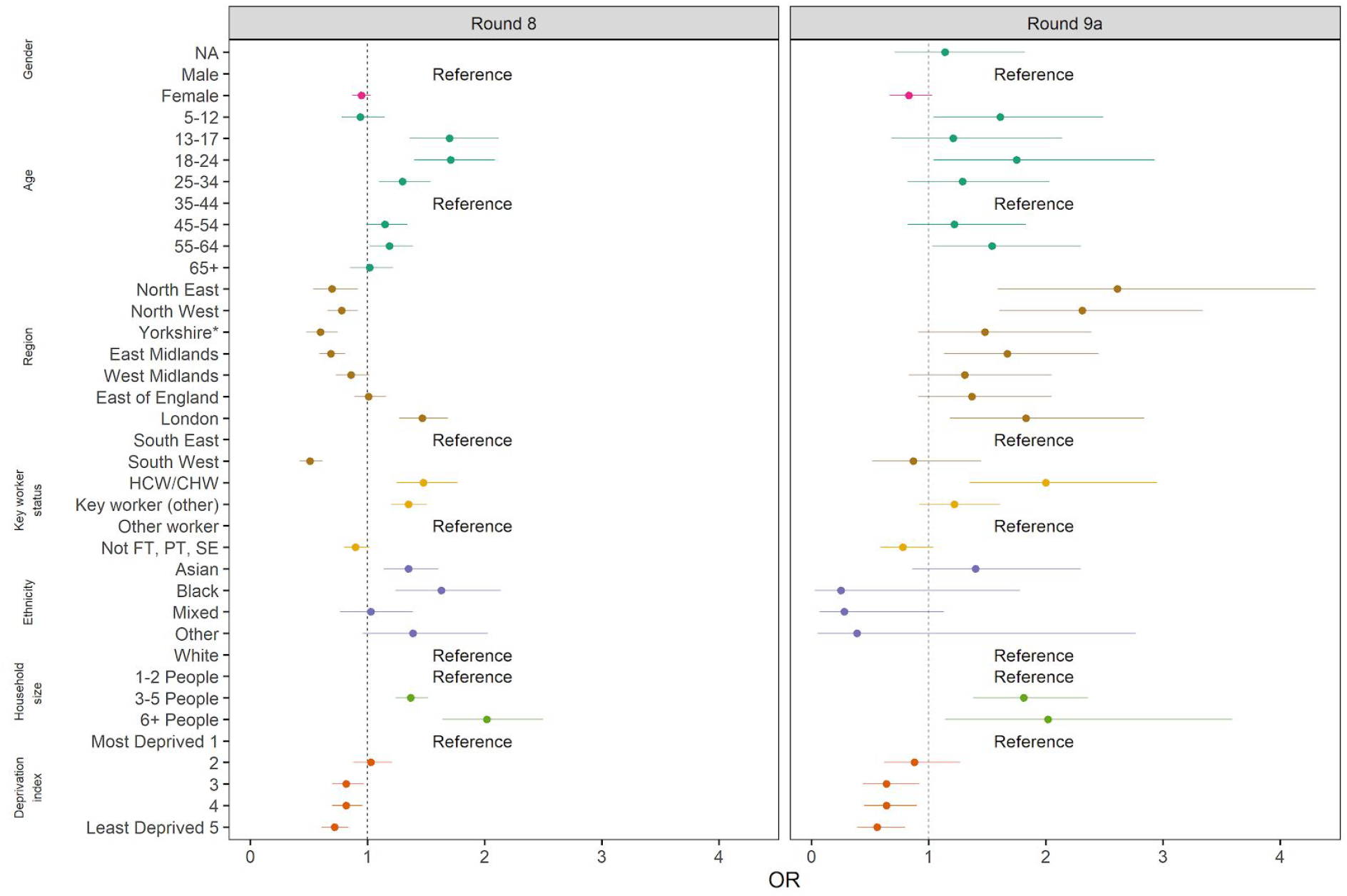
Estimated odds ratios and 95% confidence intervals for mutually-adjusted logistic regression model of swab-positivity for round 8 and round 9a. Models were adjusted for gender, age group, region, key worker status, ethnicity, household size, and deprivation index. The deprivation index is based on the Index of Multiple Deprivation (2019) at lower super output area. Here we group scores into quintiles, where 1 = most deprived and 5 = least deprived. HCW/CHW = healthcare or care home workers; Not FT, PT, SE = Not full-time, part-time, or self-employed. *Yorkshire and The Humber.

## Discussion

Round 9a of our study started four weeks into the third national lockdown in England, following extremely high rates of infection during December and early January 2021 [5]. These infection rates led to the highest levels of hospital admissions and daily deaths in England since the start of the pandemic. Our findings during round 8 of REACT 1 (mainly 6th to 21st January 2021) indicated an initial plateau at the start of lockdown, followed by a decline toward the second half of that round; there was a similar pattern for the proportion of those testing positive through symptomatic testing (Pillar 1 and 2) [6] in England [10]. Here we describe a marked decline in prevalence since then in all age groups and most regions.

Overall, we saw a two-thirds decline in prevalence across England, but this was larger in some regions, in particular London, where prevalence reached almost 3% overall and over 4% in younger ages during January 2021 [10]. In London, South East and West Midlands, prevalence fell by around 80%, although declines were smaller in the northern regions.

Since December 2020, the UK government has rolled out a vaccination programme to categories of individuals considered the most at risk for severe infection and death [12]. These categories are mainly age-based, with nearly all people of 70 years and over being offered the vaccine during January and February 2021. However the fall in prevalence was similar among those aged 65 years and over compared with other age groups, suggesting that if vaccines are effective at reducing transmission as well as disease, this effect is not yet a major driver of prevalence trends. Therefore, the observed falls described here are most likely due to reduced social interactions during lockdown.

As in previous reports [1,3,4,10], we found higher prevalence of swab-positivity among: people of Asian ethnicity compared with white people, those living in the most compared with least deprived areas, and people living in large compared to single households. We also found higher prevalence among hospital and care home workers compared with other workers, suggesting an on-going risk of infection among these groups.

Our study has a number of limitations. Because participation rates in our study may vary by a range of socio-demographic factors, it is possible that our sample is not fully representative of the base population, despite correcting for the sampling in our weighting procedure. However, unlike estimates based on symptomatic testing, we provide prevalence estimates among both symptomatic and non-symptomatic individuals from random samples of the population. Our study is therefore not subject to the biases driven by self-reporting, and health service capacity and performance present in similar data based only on tests of symptomatic individuals.

We ask individuals to provide a self-administered throat and nose swab (parent/guardian for children ages 5 to 12 years) which may be less reliable than a swab administered by a health professional. However, we provide detailed instructions including video instructions, and have utilised the same approach across all rounds of REACT-1, so that within-study comparisons and trends in prevalence over time should be robust. In addition, we have established a cold chain from home to laboratory to preserve integrity of the samples, and use a single lab (with well-defined quality control procedures) to exclude between-laboratory variation.

In conclusion, we have documented marked falls in prevalence in England during lockdown from January to February 2021. However, it should be noted that prevalence still remains high (prevalence now at a level last seen in England in September 2020 [13]). Also, the number of COVID-19 cases in hospitals is higher than at the peak of the first wave in April 2020. The effects of easing of social distancing when we transition out of lockdown need to be closely monitored to avoid a resurgence in infections and renewed pressure on health services.

## Methods

The REACT-1 study methods are published [9]. Since the first round of data collection in May 2020, in each subsequent round in each subsequent round between 150,000 and 175,000 randomly selected individuals ages 5 years and above, in England, have provided a self-administered throat and nose swab for RT-PCR testing for SARS-CoV-2. We use the list of National Health Service patients as a sampling frame, across all 315 lower-tier local authorities in England. Data collection over 2-3 weeks per round has been undertaken at approximately monthly intervals. We ask participants to complete an online or telephone administered questionnaire providing information on socio-demographics, symptoms and health and lifestyle. We use exponential growth models to estimate time trends and reproduction number R both nationally and regionally, and a p-spline function to produce smoothed estimates over time [14]. We provide national and regional crude (unweighted) prevalence estimates of SARS-CoV-2 infection as well as weighted estimates to take account of the sampling method and differential response. We then use multivariable logistic regression to estimate odds of swab-positivity for a number of predictor variables such as employment, deprivation, ethnicity and household size. We use a neighbourhood-based spatial smoothing method to investigate geographic variation in prevalence at sub-regional level [1,14].

Statistical analyses are carried out in R [15].

Research ethics approval was obtained from the South Central-Berkshire B Research Ethics Committee (IRAS ID: 283787).

## Data Availability

https://github.com/mrc-ide/reactidd

## Data availability

Supporting data for tables and figures are available either: in this Google [spreadsheet](https://docs.google.com/spreadsheets/d/1HWsqeFdRs4g7-k0_IkA9au7zXDcn4XefdtrWETqj-Cc/edit?usp=sharing); or in the inst/extdata directory of this GitHub R package.

## Acknowledgements

SR, CAD acknowledge support: MRC Centre for Global Infectious Disease Analysis, National Institute for Health Research (NIHR) Health Protection Research Unit (HPRU), Wellcome Trust (200861/Z/16/Z, 200187/Z/15/Z), and Centres for Disease Control and Prevention (US, U01CK0005-01-02). GC is supported by an NIHR Professorship. HW acknowledges support from an NIHR Senior Investigator Award and the Wellcome Trust (205456/Z/16/Z). PE is Director of the MRC Centre for Environment and Health (MR/L01341X/1, MR/S019669/1). PE acknowledges support from Health Data Research UK (HDR UK); the NIHR Imperial Biomedical Research Centre; NIHR HPRUs in Chemical and Radiation Threats and Hazards, and Environmental Exposures and Health; the British Heart Foundation Centre for Research Excellence at Imperial College London (RE/18/4/34215); and the UK Dementia Research Institute at Imperial (MC_PC_17114). We thank The Huo Family Foundation for their support of our work on COVID-19.

We thank key collaborators on this work – Ipsos MORI: Kelly Beaver, Sam Clemens, Gary Welch, Nicholas Gilby, Kelly Ward and Kevin Pickering; Institute of Global Health Innovation at Imperial College: Gianluca Fontana, Dr Hutan Ashrafian, Sutha Satkunarajah, Didi Thompson and Lenny Naar; Molecular Diagnostic Unit, Imperial College London: Prof. Graham Taylor; North West London Pathology and Public Health England for help in calibration of the laboratory analyses; Patient Experience Research Centre at Imperial College and the REACT Public Advisory Panel; NHS Digital for access to the NHS register; and the Department of Health and Social Care for logistic support. SR acknowledges helpful discussion with attendees of meetings of the UK Government Office for Science (GO-Science) Scientific Pandemic Influenza – Modelling (SPI-M) committee.

